# Early Tocilizumab Dosing is Associated with Improved Survival In Critically Ill Patients Infected With Sars-CoV-2

**DOI:** 10.1101/2020.10.27.20211433

**Authors:** Russell M. Petrak, Nicholas W. Van Hise, Nathan C. Skorodin, Robert M. Fliegelman, Vishnu Chundi, Vishal Didwania, Alice Han, Brian P. Harting, David W. Hines

**Author notes:** Corresponding Author: Russell M. Petrak M.D., 901 McClintock Dr., Burr Ridge, IL 60527. Alternate Corresponding Author: Nathan C Skorodin Pharm.D., 901 McClintock Dr., Burr Ridge, IL 60527.

## Abstract

**Background:** SARS-CoV-2 is a novel coronavirus that has rapidly expanded to become a pandemic, resulting in millions of deaths worldwide. The cytokine storm is caused by the release of inflammatory agents and results in a physiologic disruption. Tocilizumab is an IL-6 receptor antagonist with the ability to suppress the cytokine storm in critically ill patients infected with SARS-CoV-2.

**Methods:** This was a multi-center study of patients infected with SARS-CoV-2, admitted between 3/13/20 and 4/16/20, requiring mechanical ventilation. Parameters that were evaluated included age, sex, race, usage of steroids, inflammatory markers, and comorbidities. Early dosing was defined as a tocilizumab dose administered prior to or within one (1) day of intubation. Late dosing was defined as a dose administered greater than one (1) day after intubation. A control group that was treated only with standard of care, and without tocilizumab, was utilized for comparison (untreated).

**Findings:** We studied 118 patients who required mechanical ventilation. Eighty-one (81) received tocilizumab, compared to 37 who were untreated. Early tocilizumab therapy was associated with a statistically significant decrease in mortality as compared to patients who were untreated (p=0.003). Dosing tocilizumab late was associated with an increased mortality compared to the untreated group (p=0.006).

**Interpretation:** Early tocilizumab administration was associated with decreased mortality in critically ill SARS-Co-V-2 patients, but a potential detriment was suggested by dosing later in a patient’s course.

**Funding:** This work did not receive outside funding or sponsorship.

## Introduction

SARS-CoV-2 is a novel coronavirus, first identified in Wuhan, China in late 2019^1^, that has rapidly expanded to become a worldwide pandemic. The spectrum of symptomatic COVID – 19 disease ranges from mild, nonspecific influenza-like symptoms, to a second stage characterized by progressive hypoxia and pulmonary infiltrates. The most severe stage of the illness, characterized by the production of multiple cytokines, results in end organ dysfunction, hypercoagulability and increased mortality. While no specific therapy has been identified and validated in clinical trials, tocilizumab has been suggested as a possible immunomodulatory agent to offset what has been referred to as the cytokine storm^2^.

Numerous studies have reviewed the utility of tocilizumab with varying results^2–7^.This study was designed to identify the utility and timing of tocilizumab dosing for critically ill patients infected with SARS-CoV-2.

## Methods

This study was conducted by Metro Infectious Disease Consultants (MIDC), a fully integrated infectious disease (ID) private practice composed of 108 ID physicians. This was a multi-center study evaluating patients requiring mechanical ventilation for a SARS-CoV-2 infection who were admitted between 3/13/20 and 4/16/20. Patients were evaluated retrospectively and concurrently through chart review and direct interactions with the prescribing MIDC physician. This study was independently approved by Western IRB. Tocilizumab was prescribed at the discretion of the ID physician with consideration of local hospital treatment protocols. Parameters for evaluation included the patient’s age, sex, race, date of admission, usage of steroids, hydroxychloroquine (HCQ) and azithromycin (AZ) in combination, and remdesivir therapy. Comorbidities evaluated included age greater than sixty (60) years old, diabetes, chronic obstructive pulmonary disease, bronchospastic illness, chronic cardiac or renal disease, immunodeficiency or neoplastic disease. The tocilizumab dose was recorded by the timing of administration in relation to the date of hospital admission and mechanical ventilation (MV). Early dosing was defined as a tocilizumab dose administered prior to or within one (1) day of intubation. Late dosing was defined as a dose administered greater than one (1) day after intubation. Markers of inflammation including ferritin, D-dimer, LDH, and CRP were recorded at the time of dosing for the treatment group and time of intubation for the control group. Patient dispositions were recorded as discharged from the hospital or expired.

### Statistical Analysis

Descriptive statistics for baseline patient characteristics, clinical variables, and outcomes were summarized using means and standard deviations for continuous variables and counts and percentages for categorical variables. Continuous outcomes were evaluated using t-tests. Chi-squared tests or Fisher Exact tests were used to evaluate categorical outcomes. Potential confounding variables including age, gender, race, and comorbidities were adjusted for using propensity score methods. Propensity scores, defined as the probability of being in either intervention group, were calculated from a regression model with the dose timing (early vs late) as the outcome and baseline demographic and clinical variables of each participant as predictor variables. To adjust the sample for these baseline confounding variables, the propensity scores were converted to weights (“inverse probability of treatment weights”, or IPTW), and the outcomes were then compared using weighted versions of the Chi-Square test or Fisher’s exact test for categorical outcomes or a weighted version of the t-test for continuous outcomes. P-values for both the unadjusted and weighted versions of the tests are reported.

To confirm robustness of the propensity score weighted test results, multivariable regression models were also fit for each outcome, adjusting for baseline patient demographic variables. Statistical significance for all methods was defined as a p-value ≤ 0.05.

## Results

One hundred eighteen (118) hospitalized patients who required mechanical ventilation were evaluated from 23 hospitals in 4 states and were treated by 25 individual MIDC physicians (Table 1). Of these, eighty-one (81) patients received tocilizumab with 37 treated early and 44 late. An additional 37 control patients were treated with standard of care only, excluding tocilizumab therapy (untreated).

**Table 1.** Baseline Demographics and Clinical Characteristics - Tocilizumab vs No Tocilizumab.

| Variable | Label | Total<br>(N=118) | Tocilizumab:<br>No (N=37) | Tocilizumab:<br>Yes (N=81) | Unadjusted<br>p-value | PS-Weighted<br>p-value |
| --- | --- | --- | --- | --- | --- | --- |
| Age |  | 58.24<br>(12.98) | 62.33<br>(12.89) | 56.32<br>(12.65) | 0.019 | 0.608 |
| Gender |  |  |  |  | 0.406 | 0.457 |
|  | Female | 43<br>(36.44%) | 16 (43.24%) | 27 (33.33%) |  |  |
|  | Male | 75<br>(63.56%) | 21 (56.76%) | 54 (66.67%) |  |  |
| Race |  |  |  |  | > 0.999 | 0.902 |
|  | Non-White | 73<br>(61.86%) | 23 (62.16%) | 50 (61.73%) |  |  |
|  | White | 45<br>(38.14%) | 14 (37.84%) | 31 (38.27%) |  |  |
| Comorbidities Present |  | 82<br>(69.49%) | 33 (89.19%) | 49 (60.49%) | 0.003 | 0.357 |
| Ferritin |  | 2049.15<br>(2156.77) | 2001.93<br>(1818.25) | 2072.1<br>(2315.15) | 0.875 | 0.968 |
| D-dimer |  | 1207.31<br>(7073.24) | 3679.52<br>(12131.76) | 8.66 (9.16) | 0.015 | 0.07 |
| C-reactive Protein |  | 67.2<br>(100.53) | 95.62<br>(106.79) | 55.02<br>(96.04) | 0.079 | 0.517 |
| Steroid Usage |  | 76<br>(64.41%) | 26 (70.27%) | 50 (61.73%) | 0.489 | 0.433 |
| Hydroxychloroquine + Azithromycin |  | 116<br>(98.31%) | 37 (100%) | 79 (97.53%) | > 0.999 | 0.361 |
| Remdesivir |  | 3 (2.54%) | 2 (5.41%) | 1 (1.23%) | 0.231 | 0.547 |

The average age for all patients was 58.2 years, 62.3 for untreated patients, and 56.3 for those treated with tocilizumab. Seventy-five (63.6%) of the patients were male. Forty-five (38%) patients were White and 73 patients (62%) were Non-white. Comorbidities were present in 82 (69.5%) patients. Seventy-six (64.4%) patients received steroid therapy, and 116 (98.3%) received HCQ and AZ in combination at standard doses. Inflammatory markers were elevated in all patients. Table 1 shows the baseline demographics and clinical characteristics of all the patients who were included in the study, as well as the comparisons of those who received tocilizumab and those that were untreated. Both the unadjusted and propensity score weighted results show no significant differences in any baseline demographic or clinical characteristics between patients who received tocilizumab and patients who did not.

### Outcomes

Overall, sixty-one patients (51.7%) expired, 40 of which received tocilizumab and 21 were untreated (p=0.285). Table 2 shows the logistic regression model with mortality as the outcome and tocilizumab therapy, steroid use, and demographics as predictor variables. These results confirm the propensity score weighted results, showing no significant difference in the odds of mortality for all patients receiving tocilizumab regardless of timing compared to patients who were untreated, holding all other covariates constant (OR [95% CI] = 0.828 [0.34, 1.98], p = 0.670).

**Table 2.** Logistic Regression with Mortality as the Outcome - Tocilizumab vs No Tocilizumab.

| Coefficient | Odds Ratio | 95% CI | t-value | p-value |
| --- | --- | --- | --- | --- |
| Tocilizumab Given | 0.828 | (0.34, 1.98) | -0.426 | 0.670 |
| Age | 1.044 | (1.01, 1.08) | 2.226 | 0.026 |
| Gender (Male) | 1.435 | (0.65, 3.2) | 0.893 | 0.372 |
| Race (Non-White) | 2.432 | (1.08, 5.66) | 2.114 | 0.035 |
| Comorbidities Present | 0.680 | (0.24, 1.87) | -0.743 | 0.458 |
| Steroids | 1.208 | (0.54, 2.7) | 0.463 | 0.643 |

Of the 37 patients who received tocilizumab early, six (16.2%) expired, compared to 21 (56.8%) of the 37 untreated patients (p=0.002). Table 3 shows the results of a logistic regression model with mortality as the outcome and early tocilizumab therapy, steroid use, and demographics as predictor variables. These results confirm the propensity score weighted results, as early tocilizumab therapy was associated with a statistically significant decrease in mortality as compared to patients who were untreated, holding all other covariates constant (OR [95% CI] = 0.15 [0.04, 0.50], p = 0.003).

**Table 3.** Logistic Regression with Mortality as the Outcome - Early Tocilizumab vs No Tocilizumab.

| Coefficient | Odds Ratio | 95% CI | t-value | p-value |
| --- | --- | --- | --- | --- |
| Tocilizumab Given Early | 0.150 | (0.04, 0.5) | -2.963 | 0.003 |
| Age | 1.067 | (1.01, 1.13) | 2.278 | 0.023 |
| Gender (Male) | 0.788 | (0.25, 2.44) | -0.414 | 0.679 |
| Race (Non-White) | 1.699 | (0.54, 5.72) | 0.887 | 0.375 |
| Comorbidities Present | 0.553 | (0.1, 3.16) | -0.682 | 0.495 |
| Steroids | 0.507 | (0.14, 1.71) | -1.073 | 0.283 |

Thirty-four (77.3%) of 44 patients who received tocilizumab late expired compared to 21 (56.8%) of 37 patients who were untreated (p=0.006). Table 4 shows the results of a logistic regression model with mortality as the outcome and late tocilizumab therapy, steroid use, and demographics as predictor variables. These results show the odds of mortality are 3.513 times higher for patients receiving the dose of tocilizumab late compared to patients who were untreated, holding all other covariates constant (OR [95% CI] = 3.513 [1.15, 11.97], p = 0.033).

**Table 4.** Logistic Regression with Mortality as the Outcome - Late Tocilizumab vs No Tocilizumab.

| Coefficient | Odds Ratio | 95% CI | t-value | p-value |
| --- | --- | --- | --- | --- |
| Tocilizumab Given Late | 3.513 | (1.15, 11.97) | 2.133 | 0.033 |
| Age | 1.059 | (1.01, 1.11) | 2.316 | 0.021 |
| Gender (Male) | 0.973 | (0.32, 2.82) | -0.050 | 0.960 |
| Race (Non-White) | 2.631 | (0.89, 8.26) | 1.718 | 0.086 |
| Comorbidities Present | 0.823 | (0.19, 3.55) | -0.262 | 0.793 |
| Steroids | 1.353 | (0.44, 4.1) | 0.536 | 0.592 |

## Discussion

We present a multi-center, multi-state, observational study that displays a real-time evaluation of tocilizumab usage in community hospitals for critically ill SARS-CoV-2 infected patients. Most therapeutic agents presently utilized for SARS-CoV-2 have recommended guidelines suggesting the most efficacious timing for administration^8–9^. This timing has not been defined for tocilizumab secondary to conflicting data on mortality benefits. With a paucity of clinically effective SARS-CoV-2 therapies available, the timing of a therapeutic dosing window for tocilizumab is essential. This study suggests that early tocilizumab administration is associated with a mortality benefit in a critically ill SARS-CoV-2 infected patient population.

Multiple reports have previously reviewed the impact of tocilizumab therapy on mortality. Results have varied significantly with several publications failing to show a mortality benefit. These studies suffer from a retrospective design, a potential type 2 error, and/or lack of a control group. Minimal data comparing the timing of tocilizumab dosing to mechanical ventilation is available. A randomized, placebo-controlled trial not yet published showed no statistically significant mortality benefit^5^. The timing of tocilizumab was not reported, but addressed in their discussion. Fewer treatment failures (progression to mechanical ventilation, ICU admission, or death) occurred in the tocilizumab treated group than patients who received placebo. This suggests that the timing of tocilizumab administration is critical to a patient’s outcome. In contrast, several trials have shown a positive clinical impact when tocilizumab is given prior to intubation, or when oxygenation requirements markedly increase. A randomized, placebo-controlled trial, yet to be released, claims that tocilizumab reduces the need for mechanical ventilation in SARS-CoV-2 infected patients^10^.

Mortality rates for patients requiring mechanical ventilation are significant and have varied from 24.5%-79%^11–12^. Overall, 51.7% of the patients in our study expired. A statistically significant mortality differential was characterized by the timing of the tocilizumab administration. Patients who received an early tocilizumab dose showed a statistically significant decrease in mortality as compared to patients who were untreated (16.22% vs. 56.76%, p < 0.001), after adjusting for demographic characteristics and steroid use. As has been delineated in other studies, non-white and elderly patients were more likely to succumb to their illness. Although we did not identify any secondary infections in our patient population, several authors have reported an increase with tocilizumab therapy^13^.

A statistically significant increase in mortality was associated with tocilizumab administration later than one day after intubation (p=0.006). This suggests that the administration of tocilizumab is not only ineffective, but potentially deleterious, if given later in a critically ill patient’s course. The etiology of this potentially enhanced decompensation is unclear. Explanations for this outcome include selection bias of a more critically ill population, higher levels of IL-6^14^, or an inability to diminish the cytokine storm due to a delay in treatment. The results of this relatively small series should be interpreted cautiously, as further randomized studies will be needed to define this predisposition.

With a disease process that is difficult to control and predict, timing of effective therapies is essential. A delay in tocilizumab therapy is associated with a statistically significant increase in mortality regardless of sex, age, race, steroid therapy, or the presence of comorbidities. Accordingly, in an effort to optimize favorable outcomes, hospitals will need to identify barriers to early dosing. While the etiology of delay for individual patients is difficult to define, several have been identified. These include lack of drug availability, restrictive protocol requirements, or lack of consensus between consulting physicians. This data should hopefully stimulate our clinical and administrative colleagues to work collaboratively to reduce these obstructions to care.

Our study has certain limitations. This study was observational and we did not formally evaluate patients for hepatitis B or latent tuberculosis. Although no baseline characteristics were statistically significant between our groups, patients who received tocilizumab were also treated with antibiotics, anticoagulants, and steroid therapy in varying combinations. Accordingly, it is difficult to identify the most clinically influential modality or optimal therapeutic combination.

To date, no prospective randomized studies analyzing the combination of all potential COVID treatment modalities within a specific therapeutic window have been published. Pending the availability of this data, our study provides guidance to clinicians striving to identify the most efficacious timing for tocilizumab therapy. The presented data supports a mortality benefit of early tocilizumab therapy, within 1 day of intubation, and a possible detriment to later dosing. We strongly encourage the use of tocilizumab earlier in the COVID-19 treatment spectrum.

## Data Availability

A deidentified dataset and the statistical analysis will be made available to the editor in chief or delegate for a period of 30 days from the time of submission for validation of results.

